## Extended Data Figures for "Cytotoxic CD8^+^ T cells target citrullinated antigens in rheumatoid arthritis"

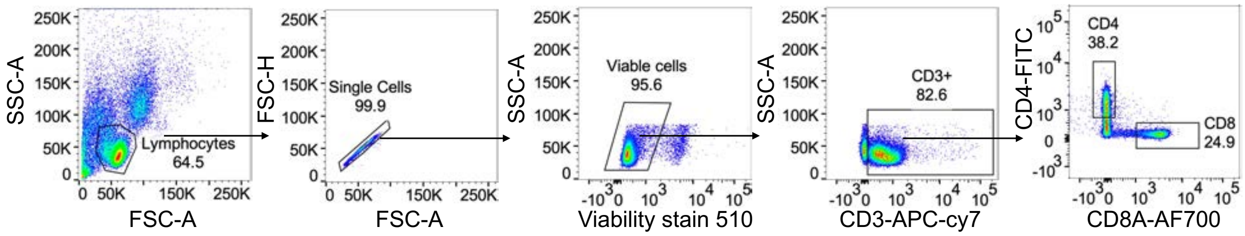


**Extended Data Fig. 1: Flow cytometric analysis of CD8$^+$ T cells in ACPA$^+$ RA patient blood.** Representative data from ACPA$^+$ RA PBMCs are displayed as flow cytometric plots, and the gating parameters for the CD8$^+$ T cell population indicated.

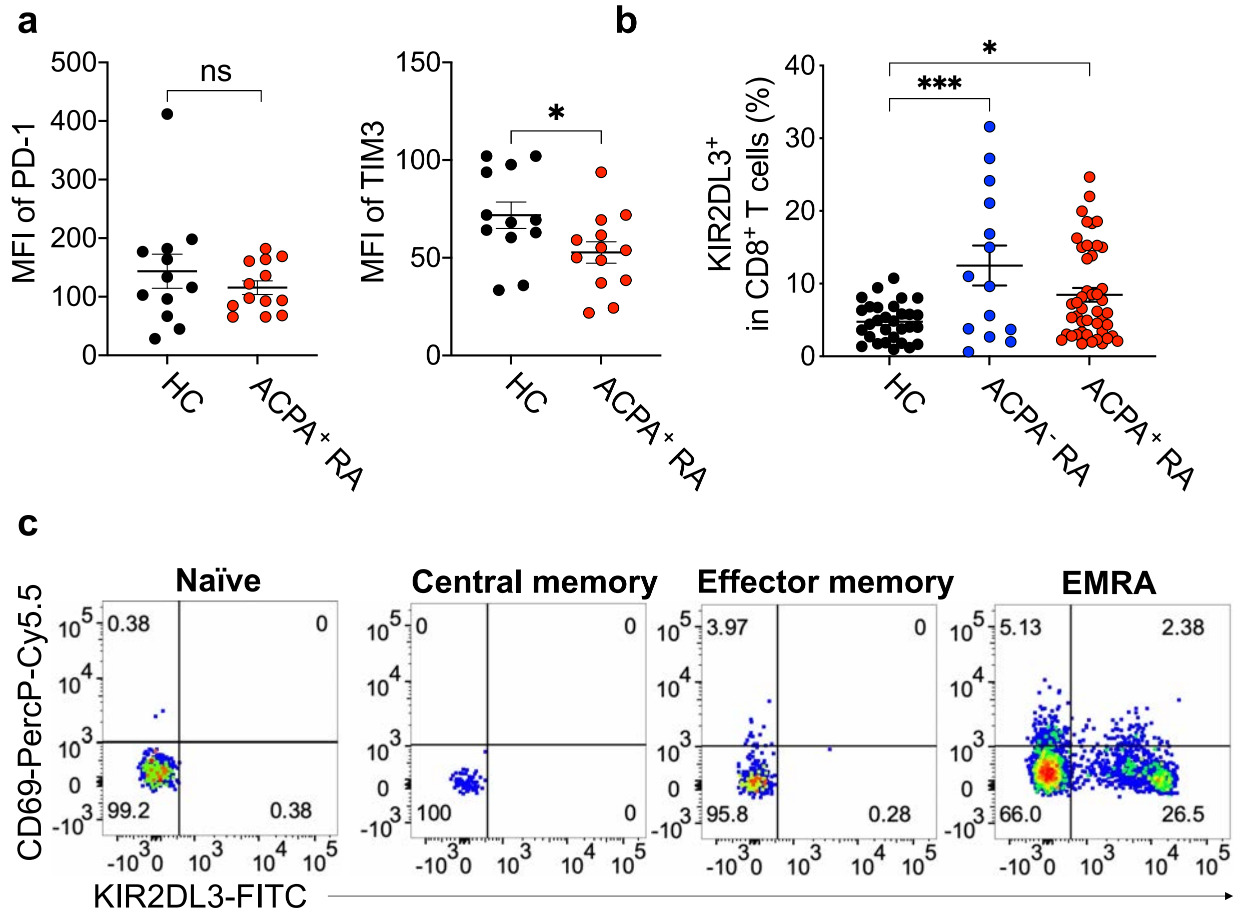


**Extended Data Fig. 3: Phenotypic characterization of CD8⁺ T cells in ACPA⁺ RA patient blood. a**, Expression of PD-1 or TIM3 in CD8⁺ T cells from PBMCs from healthy controls (*n* = 12) or ACPA⁺ RA patients (*n* = 13) measured by flow cytometry. **b**, Percentage of KIR2DL3⁺CD8⁺ T cells in HC (*n* = 30), ACPA⁻ RA (*n* = 14), or ACPA⁺ RA (*n* = 45) analyzed by flow cytometry. **c**, Representative dot plots showing expression levels of CD69 and KIR2DL3 in CD8⁺ T memory subsets (naïve, CM, EM or EMRA). Data are presented as means ± SEM. $*P < 0.05$ or $***P < 0.001$ by unpaired *t*-test (**a**) or one-way ANOVA (**b**). ns, not significant.

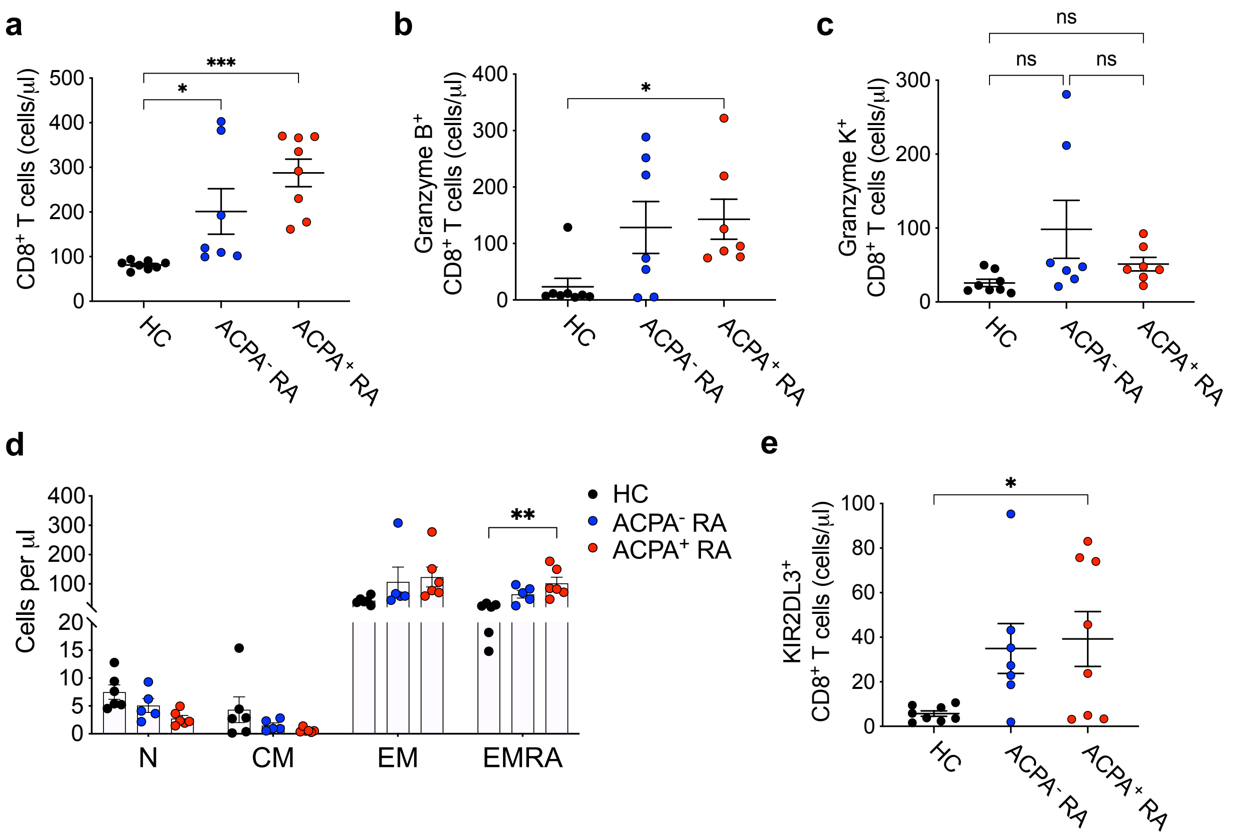


**Extended Data Fig. 3: Absolute cell counts of CD8$^+$ T cells in ACPA$^+$ RA patients.** Absolute cell numbers of total CD8$^+$ T cells (**a**), granzyme B-expressing CD8$^+$ T cells (**b**) or granzyme K (**c**)-expressing CD8$^+$ T cells, memory CD8$^+$ T cells (**d**) or KIR2DL3$^+$CD8$^+$ T cells (**e**) per 1 µl assessed by counting beads in HC (n = 6 or 8), ACPA$^-$ (n = 5 or 7) or ACPA$^+$ (n = 6, 7 or 8) RA PBMCs. Data are presented as means ± SEM. *$P < 0.05$, **$P < 0.01$ or ***$P < 0.001$ by one-way ANOVA (**a-e**). ns, not significant.

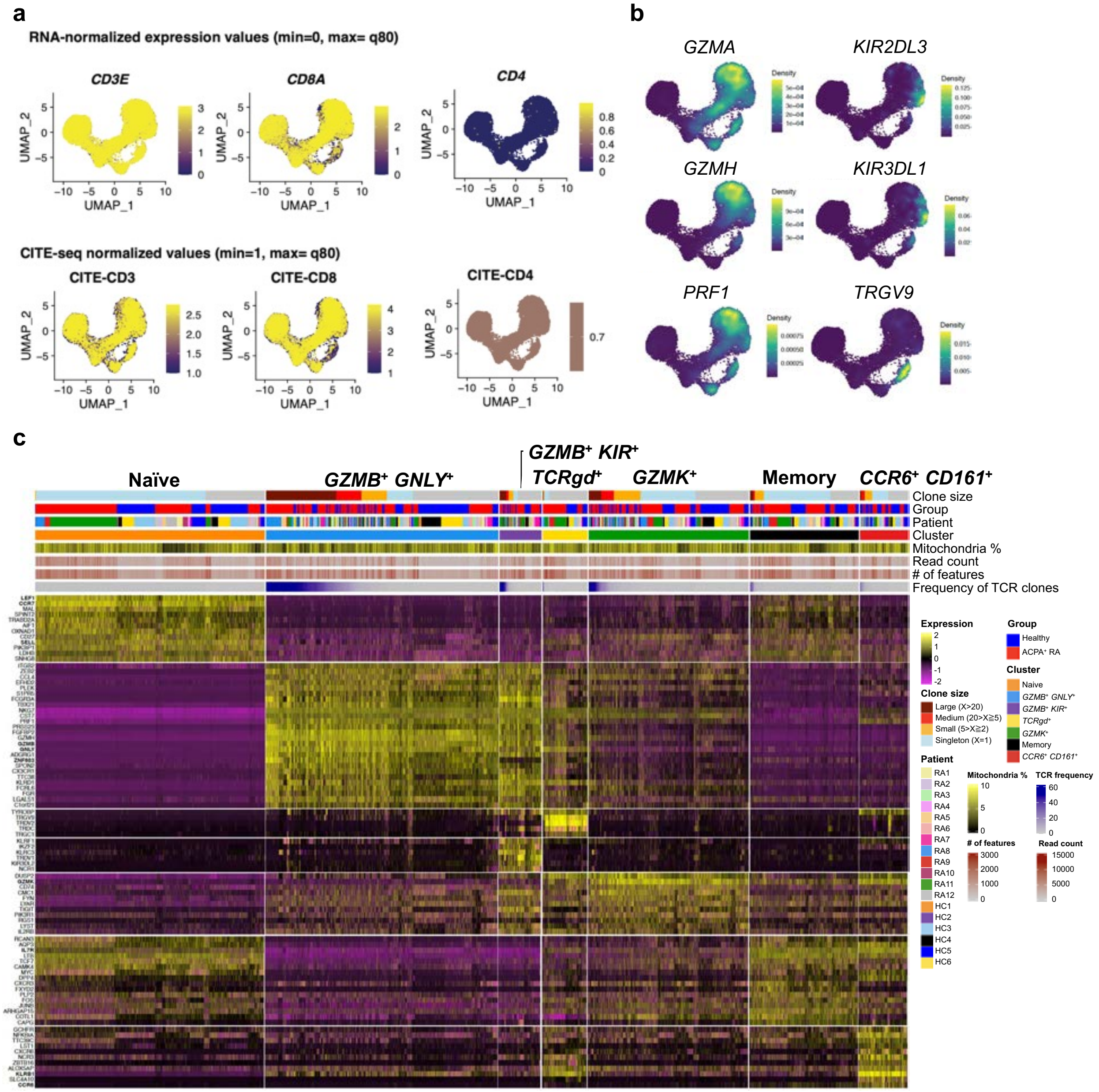


**Extended Data Fig. 4: Expression of cell type marker genes in CD8$^+$ T cells from ACPA$^+$ RA and healthy control blood. a,** mRNA and CITE-seq expression of marker genes (*CD3E*, *CD8A*, and *CD4*) used to identify and confirm analysis of CD8$^+$ T cells. Color indicates the expression level of each marker gene. **b,** Density plots display marker genes including *GZMA*, *KIR2DL3* or *TRGV9*) in CD8$^+$ T cells. **c,** Heatmap of select differentially expressed gene signatures (log2 Fold Change > 0.8) in each cluster with bars; clonal size, patients, clusters, mitochondrial gene percentage, read count, number of features in each cell and frequency of TCR clone.

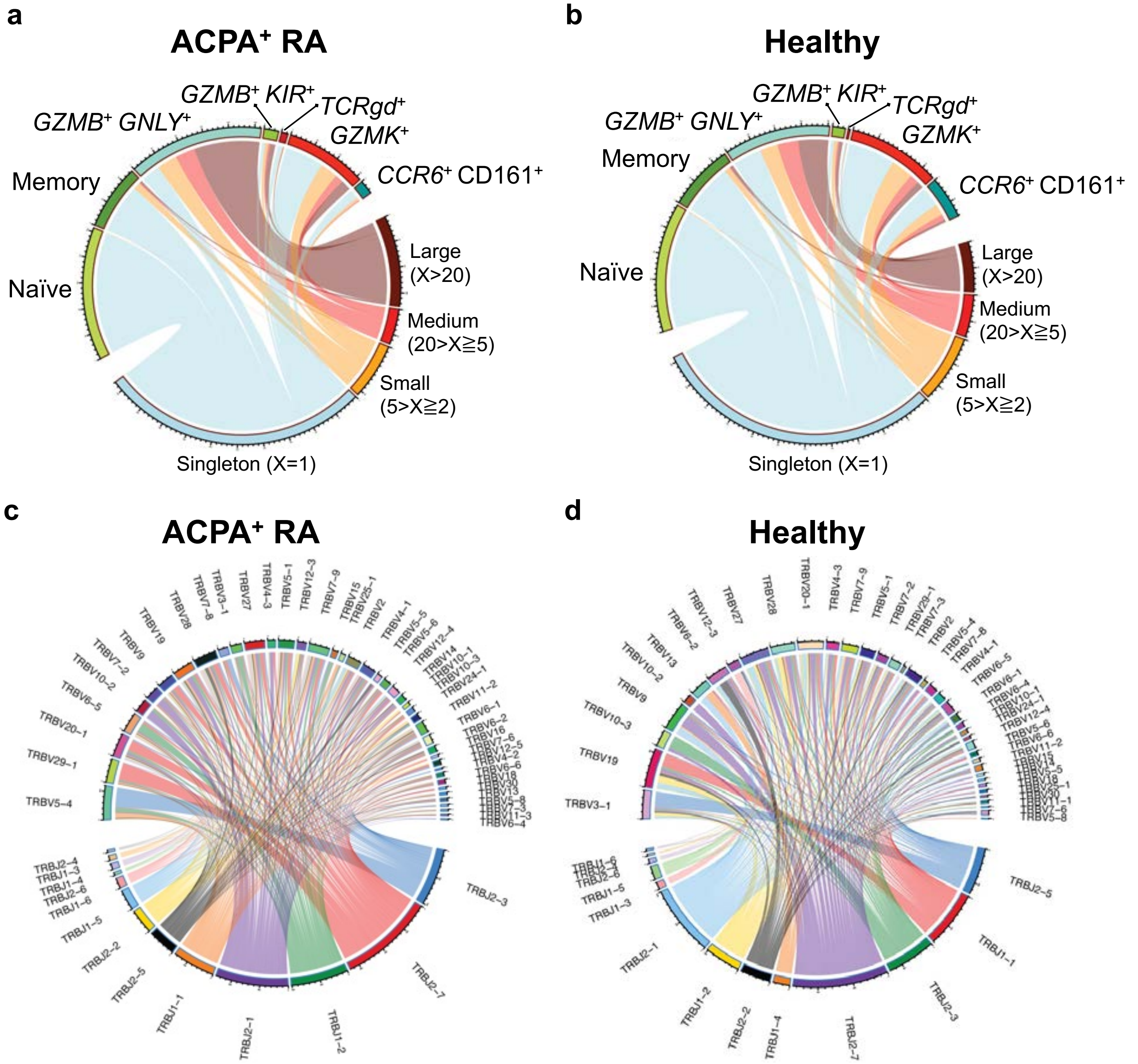


**Extended Data Fig. 5: CD8$^{+}$ T cells expressing cytotoxic mediators are clonally expanded in ACPA$^{+}$ RA blood. a,b,** Chord diagrams representing the contribution of each cluster to clonal sizes (**a**) and the shared clonotypes between clusters (**b**). **c,** TCR beta chain V- and J-gene usage of CD8$^{+}$ T cells of ACPA$^{+}$ RA blood. The width of each band represents the frequency of V- or J- gene usage.

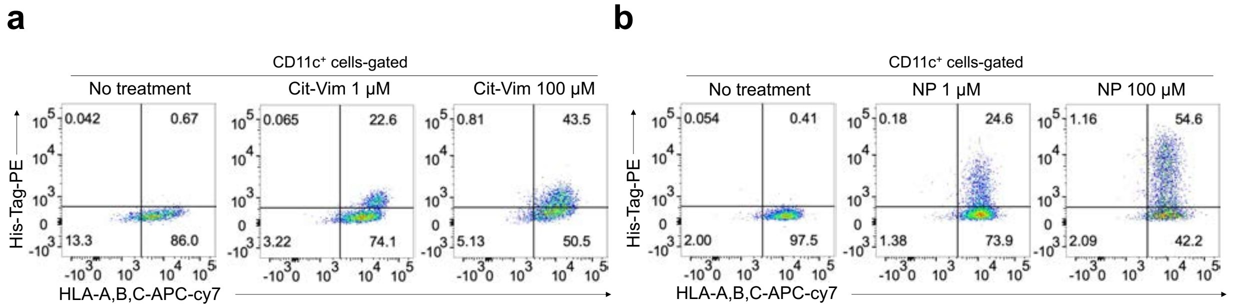


**Extended Data Fig. 6: CD11c⁺ cells from ACPA⁺ RA patients endocytose cit-vimentin. a,b,** Representative flow cytometric plots of protein endocytosis by CD11c⁺ cells in ACPA⁺ RA PBMCs show antigen-presenting cells uptake recombinant His-tagged cit-vimentin protein (**a**) or NP protein (**b**) in a d8se-dependent manner (1 or 100 μM) after 16 hr of culture.

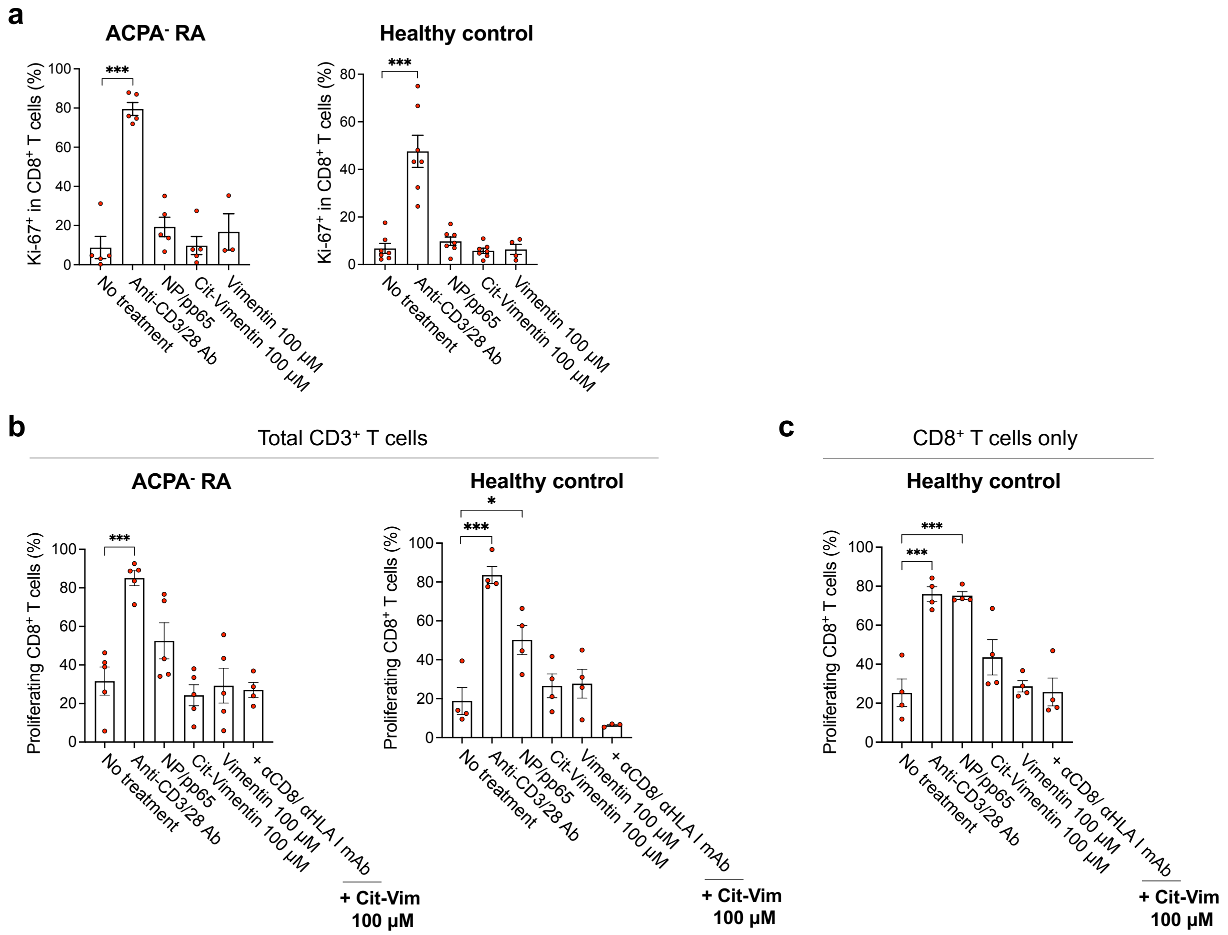


**Extended Data Fig. 7: Citrullinated vimentin does not induce the proliferation of ACPA⁻ RA or HC CD8⁺ T cells. a,** Percentage of Ki-67⁺CD8⁺ T cells in ACPA⁻ RA (*n* = 5) or HC (*n* = 7) PBMCs after stimulation with anti-CD3/28 antibodies, NP (Influenza)/ pp65 (CMV) proteins (50 µM of each), citrullinated vimentin (100 µM), or native vimentin (100 µM) for 16 hr. **b,c,** Quantification of the proliferating CD8⁺ T cells in co-culture of MoDCs with ACPA⁻ RA or HC total CD3⁺ T cells (b, n = 4 or 5) or CD8⁺ T cells only (c, n = 4). Bars represent means ± SEM. *$P < 0.05$ or ***$P < 0.001$ by ordinary one-way ANOVA with Tukey's multiple comparison test.

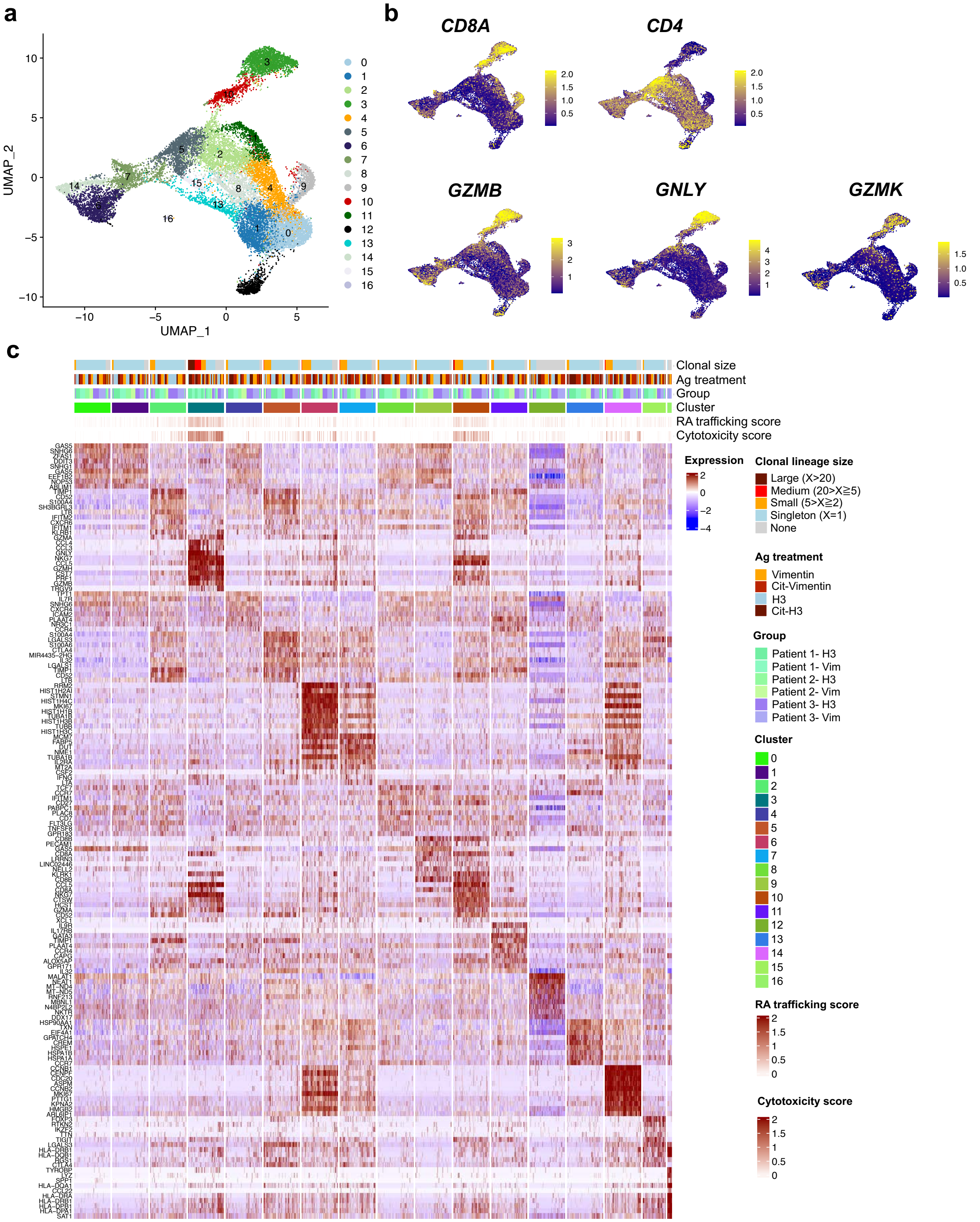


**Extended Data Fig. 8: Single-cell RNA sequencing of CD3$^+$ T cells stimulated by native or citrullinated antigens. a,** UMAP plot representing 17 distinct clusters of CD3$^+$ T cells (n = 13,354) stimulated by native or citrullinated proteins. **b,** Expression of *CD8A*, *CD4*, *GZMB*, *GNLY* or *GZMK* in the clusters. Color shows the expression level of each marker. **c,** Heatmap showing transcriptional differences in each cluster. Top bars indicate clonal size, Protein treatment, patient group, clusters and scoring scale for RA trafficking and cytotoxicity. Individual T cells are ordered by clonal size in each cluster.

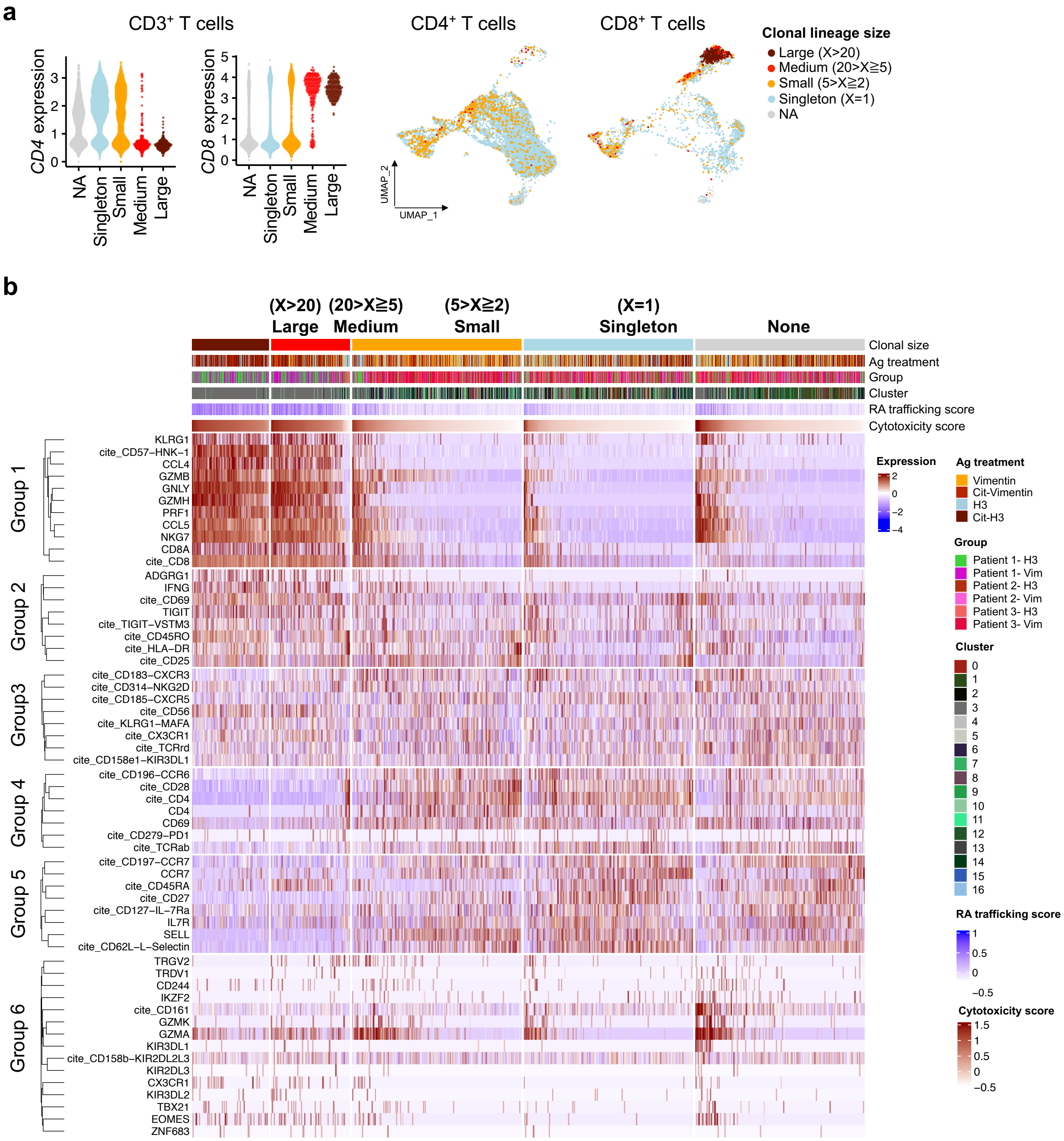


**Extended Data Fig. 9: Clonal lineages of CD4$^+$ and CD8$^+$ T cells stimulated by native or citrullinated antigens. a,** Distribution of each clonal family in *CD4*$^+$ (n = 7,885) or *CD8*$^+$ (n = 3,284) T cells. **b,** Heatmap representing 6 groups determined by unsupervised clustering based on the level of RNA and CITE-seq antibodies (cite_). Individual T cells are ordered by cytotoxicity score in each cluster.

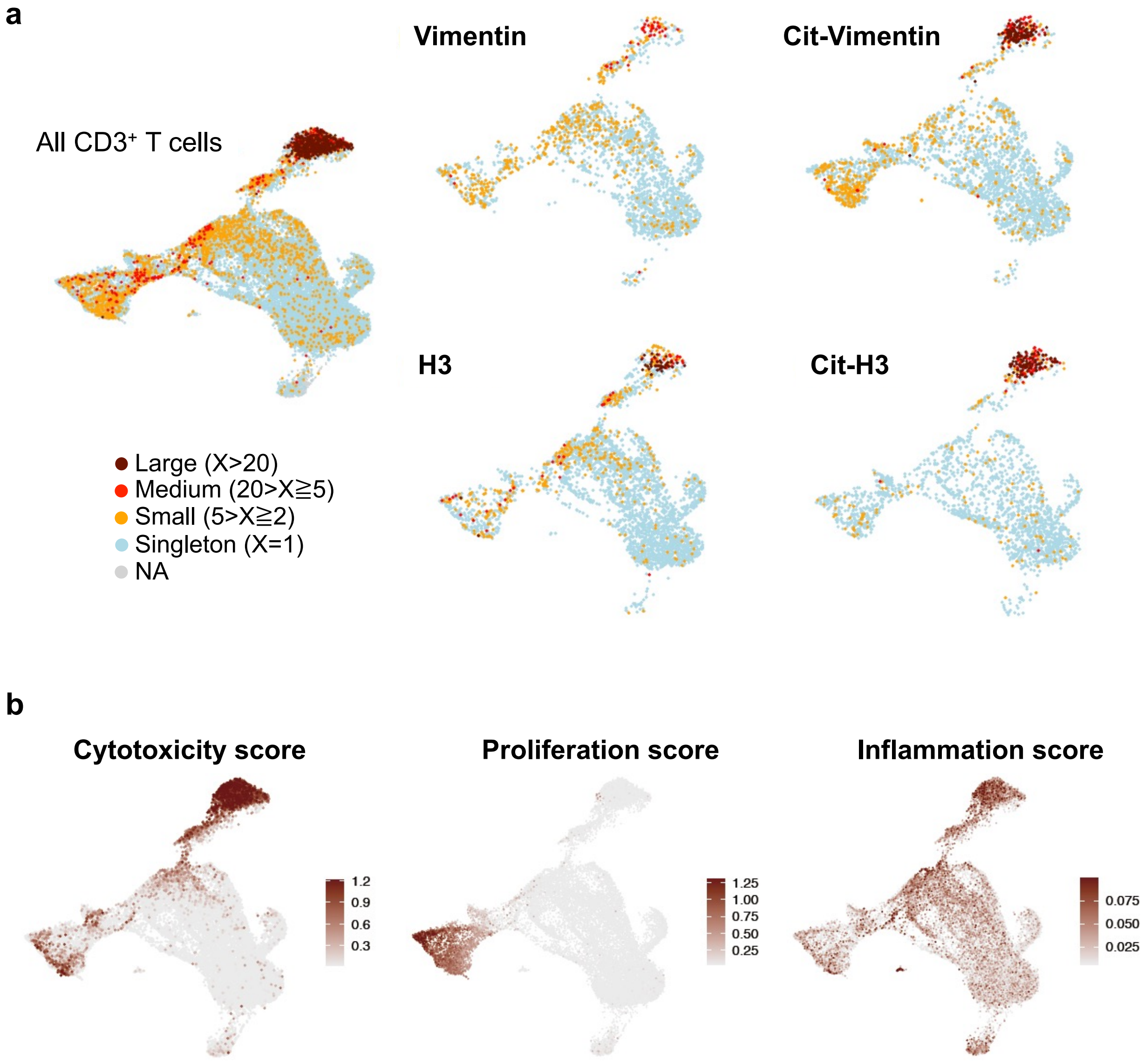


**Extended Data Fig. 10: Citrullinated antigens stimulate clonal expansion of $CD3^+$ T cells with high cytotoxicity scores. a,** UMAP plots of all $CD3^+$ T cells stimulated by native or citrullinated Vimentin or H3 integrated with TCR clonality. **b,** UMAP plots showing cytotoxicity, proliferation, or inflammation score. The score was calculated by the expression level of canonical marker genes.

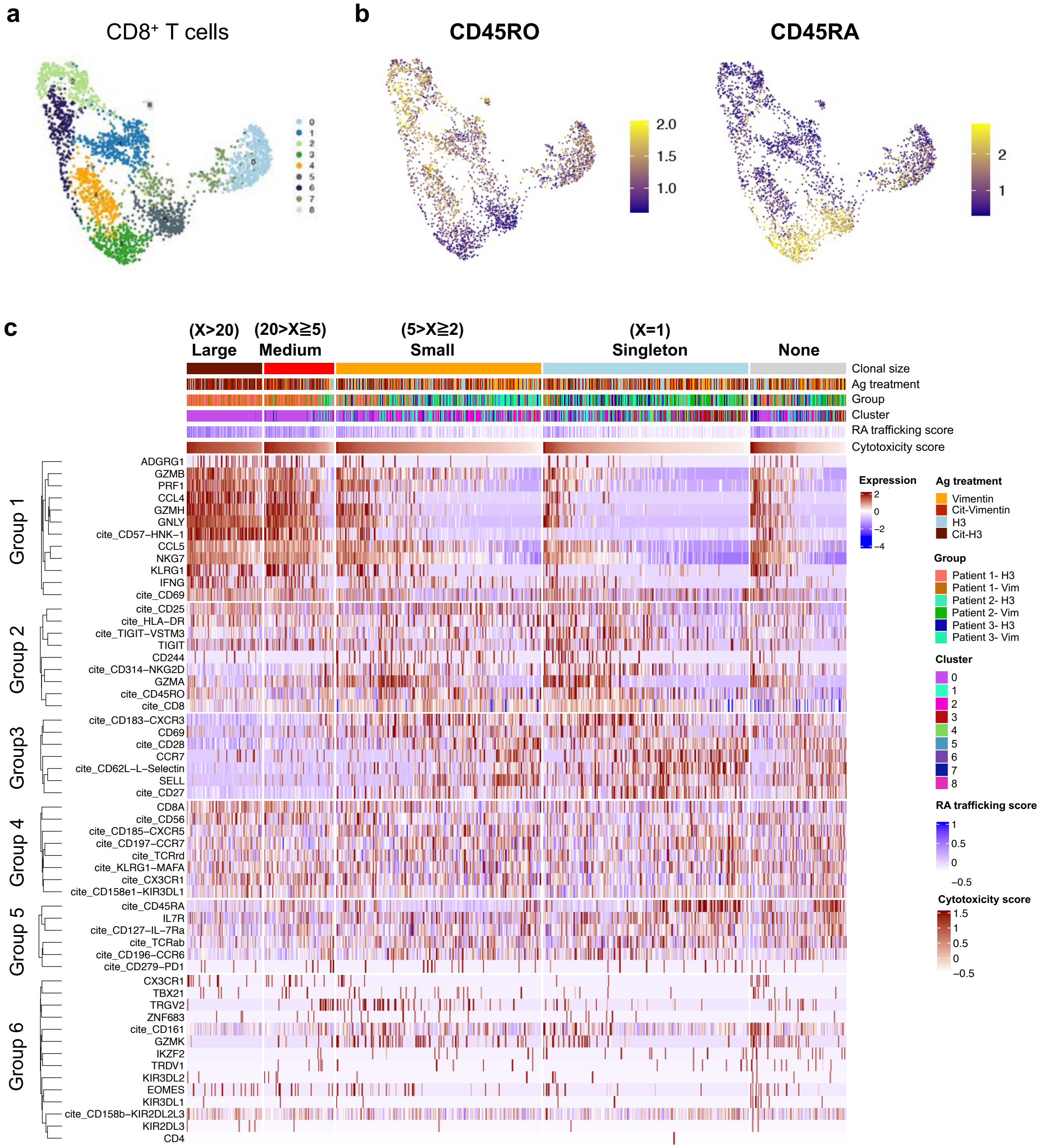


**Extended Data Fig. 11: Phenotype of CD8$^+$ T cells stimulated by native or citrullinated antigens. a,** UMAP plot of CD8$^+$ T cells having 9 transcriptionally different clusters. **b,** Expression level of CD45RO and CD45RA CITE-seq antibodies in CD8$^+$ T cells. **c,** Heatmap of 6 groups by unsupervised clustering using the level of RNA and CITE-seq antibodies (cite_).

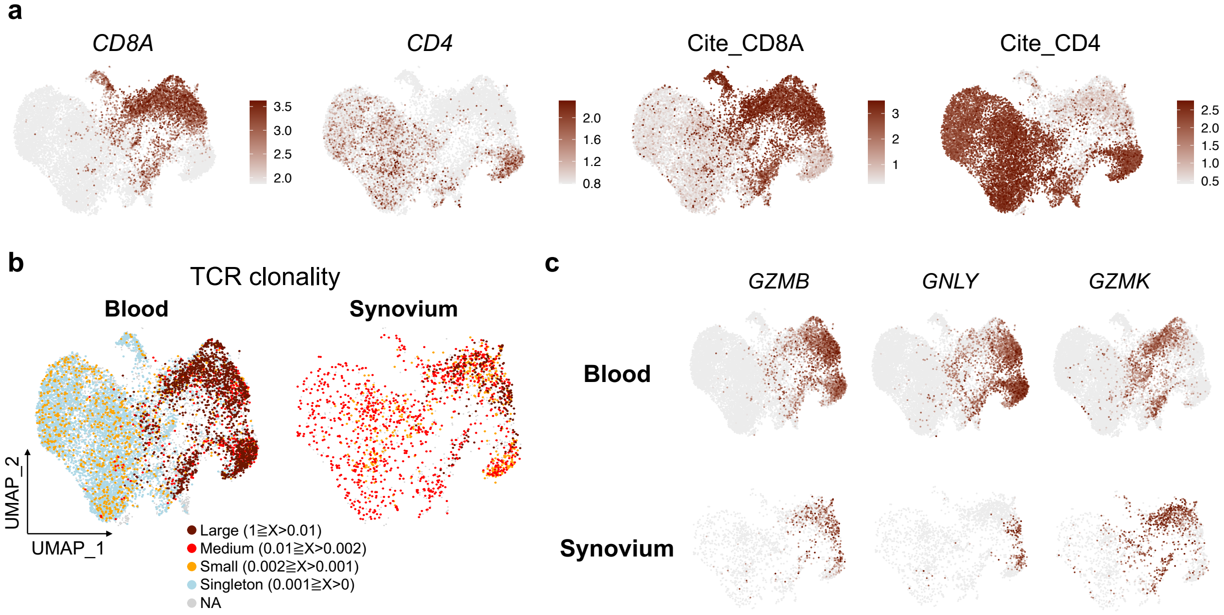


**Extended Data Fig. 12: Integrated analysis of single cell RNA-seq data from paired RA blood and synovium CD8⁺ T cells. a,** UMAP plots representing the expression levels of CD8A and CD4 in mRNA and CITE-seq Abs. **b,** UMAP plots of ACPA⁺ RA blood (*n* = 3) and synovium (n = 4) CD8⁺ T cells integrated with TCR clonality (*n* = 9,360 paired *TCRαβ* sequences). Color indicates the groups by the ratio of clonotypes in total cells. **c**, UMAP plots showing the expression of *GZMB*, *GNLY* and *GZMK* in integrated blood and synovial CD8⁺ T cells. Scale bars represent the normalized expression values.

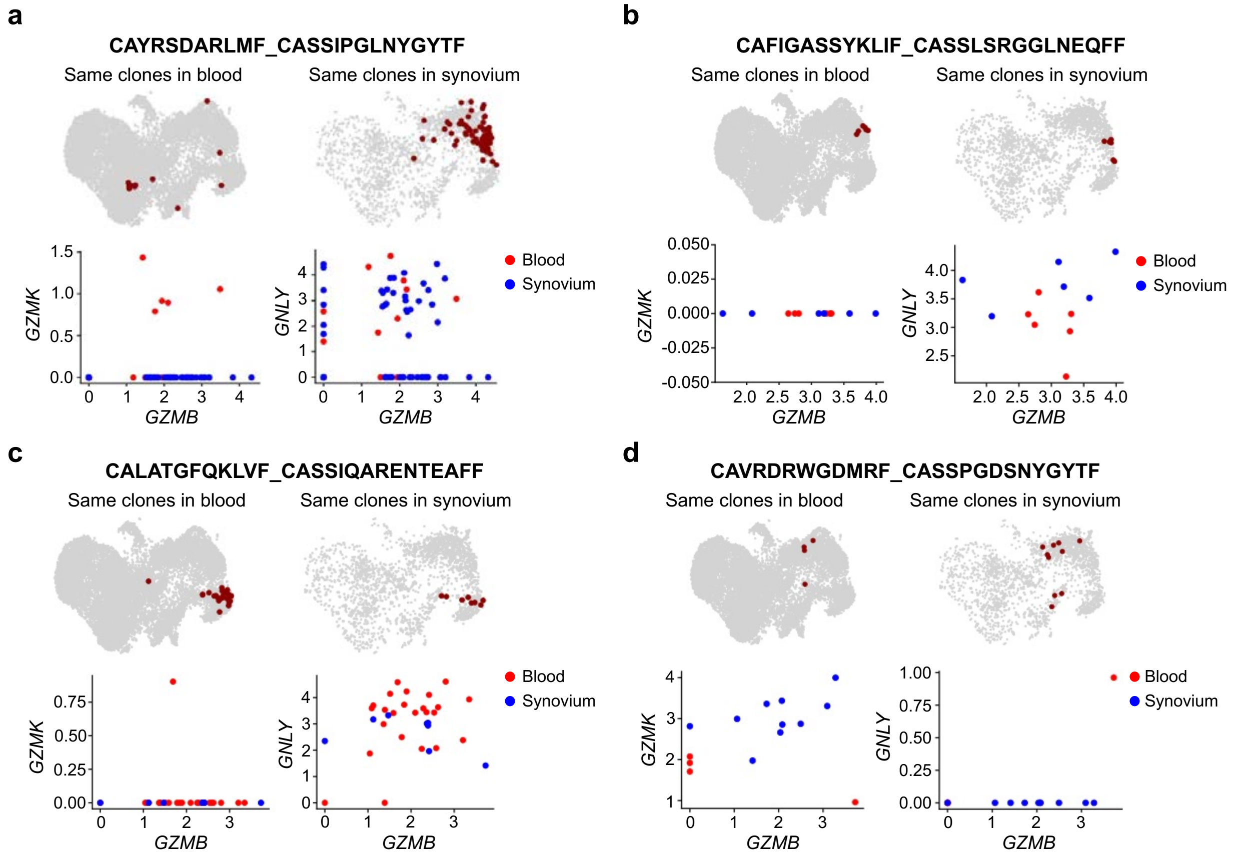


**Extended Data Fig. 13: Shared T cell clonotypes between ACPA⁺ RA blood and matched synovium. a-d,** Mapping of shared clonotypes in the matched blood and synovium from ACPA⁺ RA patients. The frequent clonotypes are represented as CDR3 alpha-beta sequences. Dot plots show the expression level of *GZMB*, *GZMK*, and/or *GNLY* in the same clonotypes located in either blood (red) and synovium (blue).

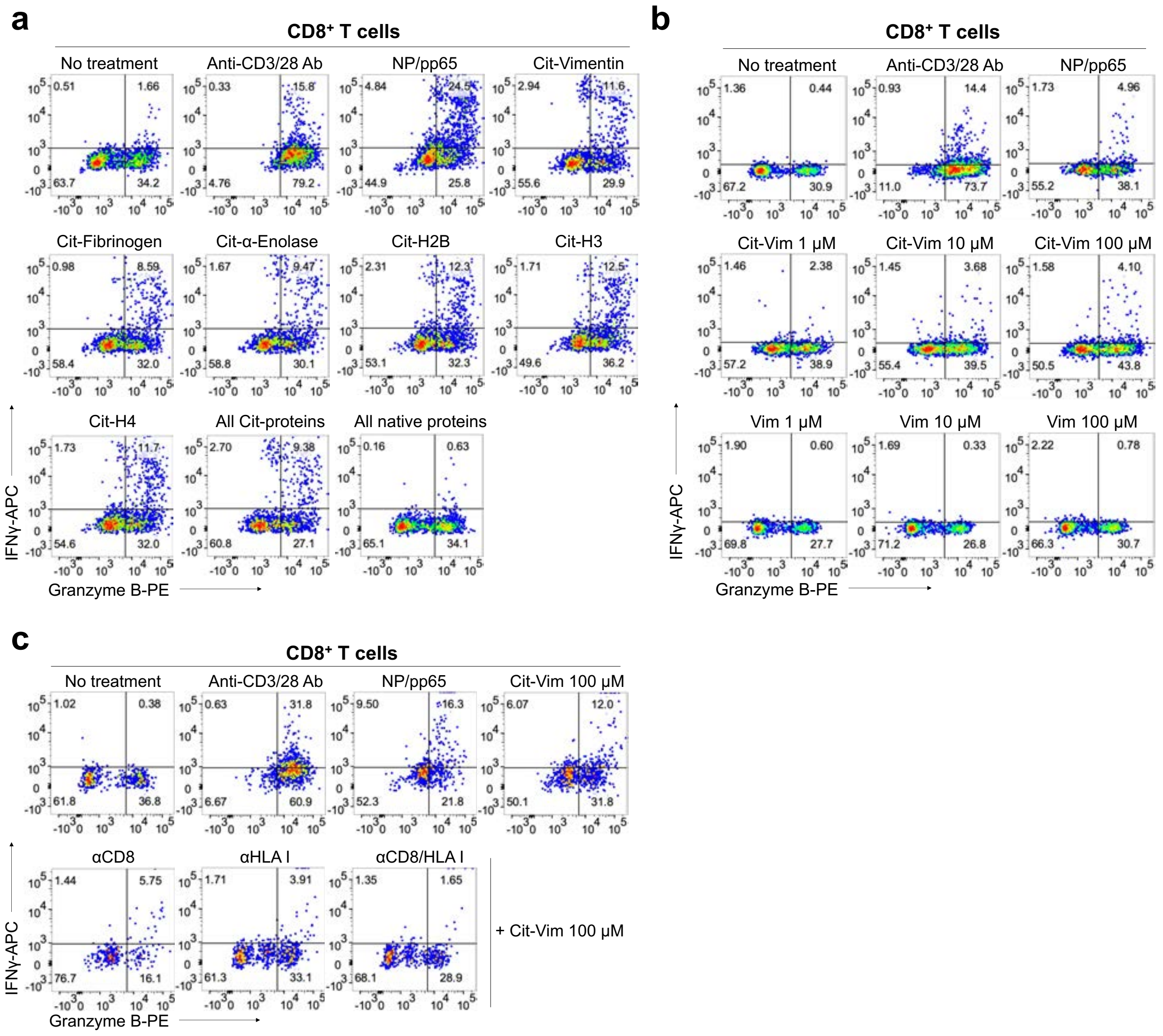


**Extended Data Fig. 14: ACPA⁺ RA patient blood CD8⁺ T cells are activated by citrullinated proteins in a CD8:HLA class I-dependent manner. a-c,** Representative flow cytometric plots of ACPA⁺ RA blood CD8⁺ T cells stimulated with citrullinated proteins (**a**), a range of concentrations of cit-vimentin or native vimentin (**b**), and in the presence of anti-CD8/ HLA class I-blocking antibodies (**c**).

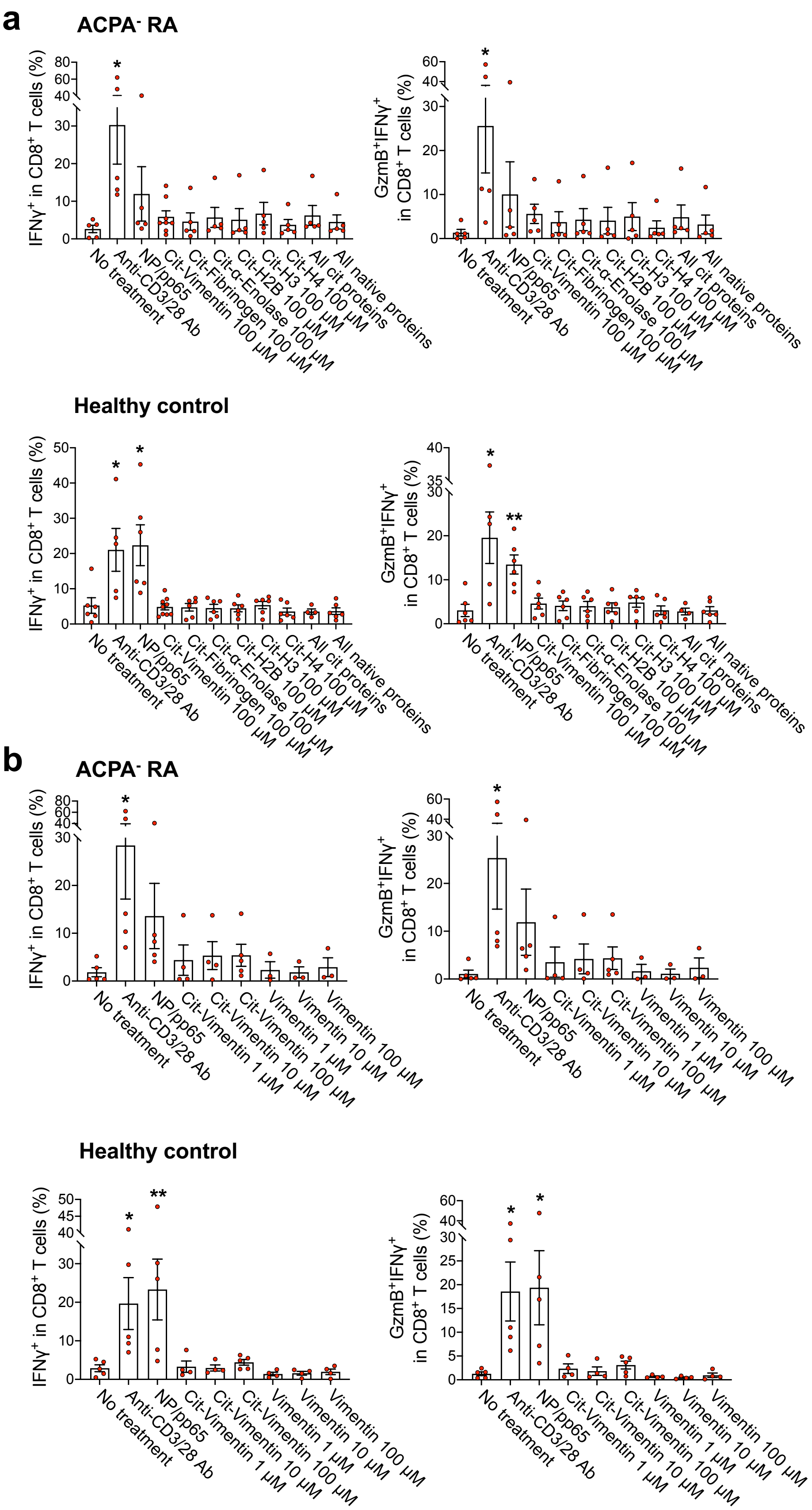


**Extended Data Fig. 15: ACPA- RA patient CD8+ T cells react with viral but not citrullinated antigens. a,** Percentages of IFNγ or GzmB-expressing CD8+ T cells from PBMCs of ACPA$^-$ RA ($n$ = 5) or HCs ($n$ = 5-6) were measured by intracellular staining after stimulation with anti-CD3/28 antibodies, NP (Influenza)/ pp65 (CMV) proteins (50 μM of each), citrullinated proteins (vimentin, fibrinogen, α-enolase, and histones H2B, H3 or H4) (100 μM), all citrullinated proteins (20 μM each), or all native proteins (20 μM each) for 16 hr. **b,** Quantification of IFNγ$^+$ or IFNγ$^+$GzmB$^+$ CD8$^+$ T cells in ACPA$^-$ RA ($n$ = 3-5) or HCs ($n$ = 4-5) PBMCs stimulated for 16 hr with cit-vimentin or native vimentin in a concentration-dependent manner. By unpaired $t$-test: *$P < 0.05$ or **$P < 0.01$ versus no treatment.

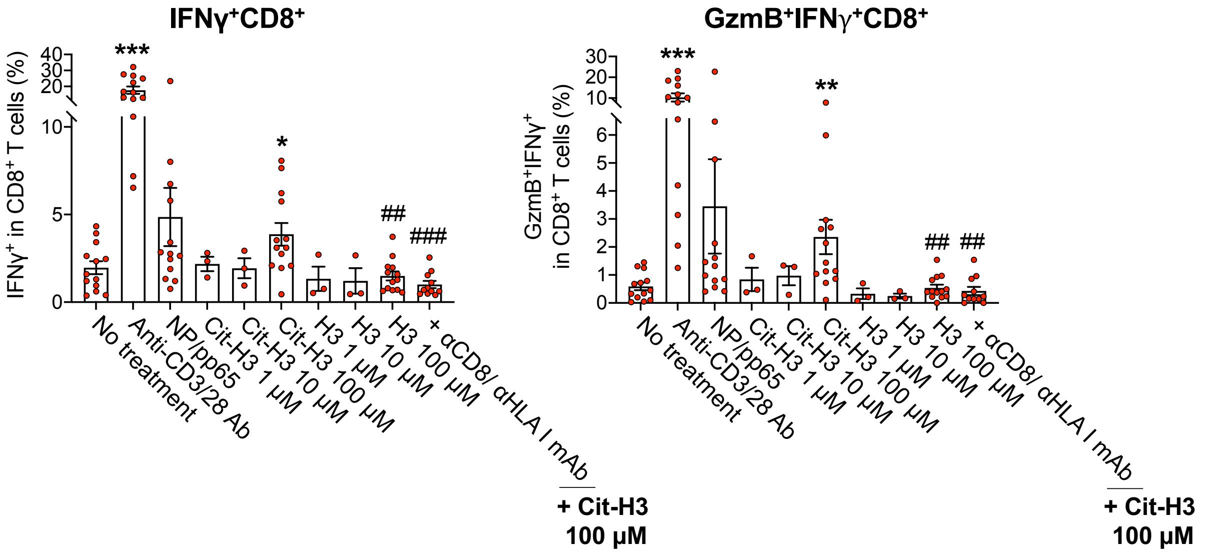


**Extended Data Fig. 16: ACPA$^+$ RA blood CD8$^+$ T cells react with cit-H3.**

Percentage of IFNγ or GzmB expressing CD8$^+$ T cells in ACPA$^+$ RA blood (n = 3-13) measured by intracellular staining after stimulation with a range of concentrations of citrullinated histone H3 (cit-H3) or native H3 protein in presence or absence of anti-CD8/HLA class I-blocking antibodies for 16 hrs. By unpaired *t*-test: $^*P < 0.05$, $^{**}P < 0.01$ or $^{***}P < 0.001$ versus no treatment, and $^{\#\#}P < 0.01$ or $^{\#\#\#}P < 0.001$ versus cit-H3 100 μM.

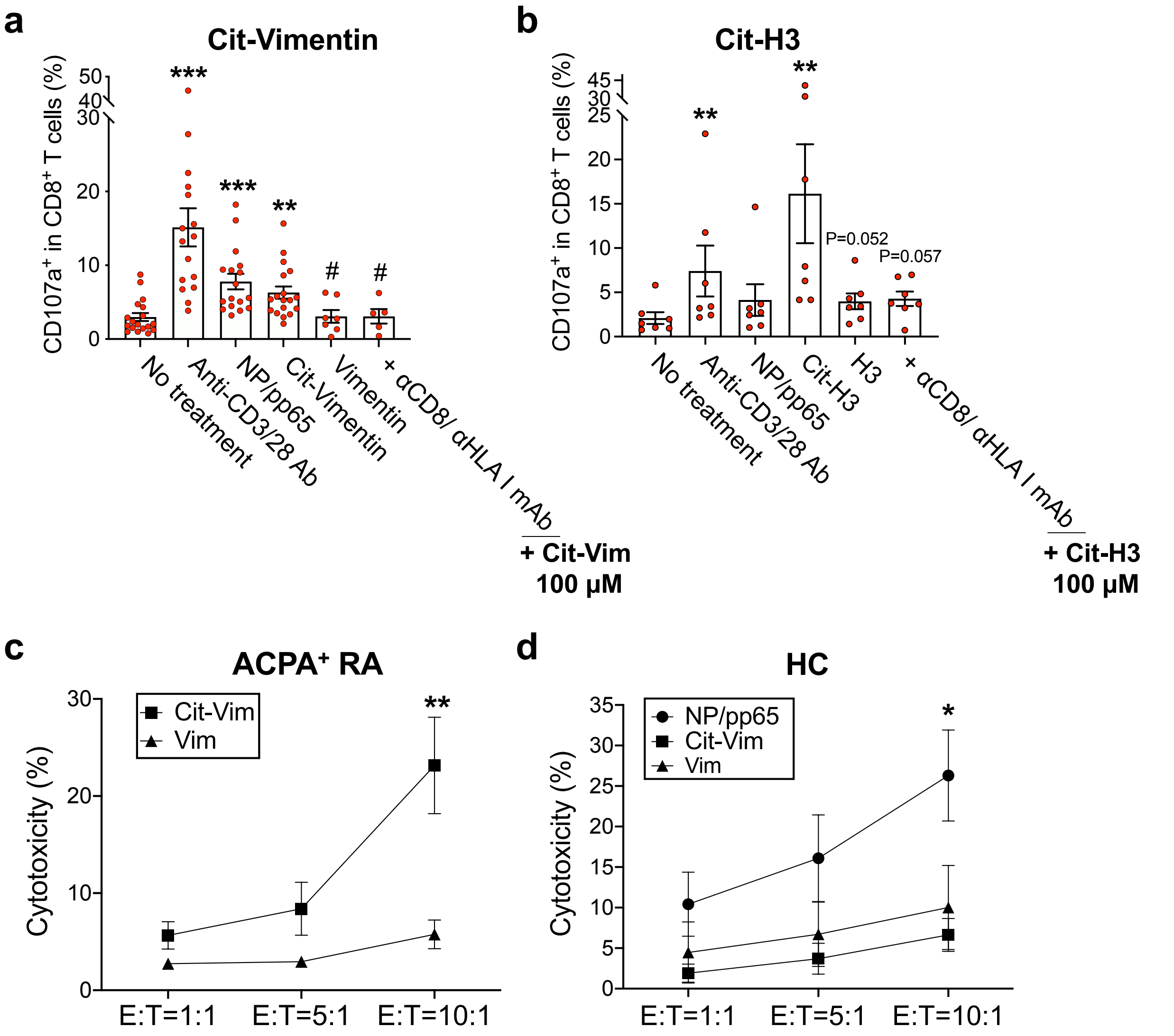


**Extended Data Fig. 17: Citrullinated antigens stimulate ACPA$^+$ RA blood CD8$^+$ T cells to exhibit cytotoxic activity and mediate cell killing. a,b,** Flow cytometry analysis of CD107a$^+$ expression by CD8$^+$ T cells was used as a measure of degranulation activity. ACPA$^+$ RA blood samples ($n$ = 5-14 (**a**), and $n$ = 7 (**b**)) were stimulated with citrullinated or native forms of the RA autoantigens vimentin and histone H3 (100 μM each), with or without anti-CD8/ HLA class I-blocking antibodies, and flow cytometry analysis performed to quantitate CD107a$^+$CD8$^+$ T cells. **c,d,** Antigen-independent cytotoxic activity of cit-vimentin, native vimentin, or viral protein stimulated CD8$^+$ T cells from ACPA$^+$ RA (**c**, $n$ = 5-7) or HC (**d**, $n$ = 3-7) blood against the DLD-1 cell line was determined using a lactate dehydrogenase (LDH) release assay. E:T ratio means effector/target cell ratio. Data are presented as means ± SEM. For **a,b**, using an unpaired $t$-test: $^{**}P < 0.01$ or $^{***}P < 0.001$ versus no treatment; $^{\#}P < 0.05$ versus cit-vimentin; P-value represented on the graph was compared with cit-vimentin. For **c,d**, using one-way ANOVA: $^{*}P < 0.05$ or $^{**}P < 0.01$.

Supplementary Tables

### Cytotoxic CD8+ T cells target citrullinated antigens in rheumatoid arthritis

**Authors:** Jae-Seung Moon[#1,2], Shady Younis[#1,2,3], Nitya S. Ramadoss[1,2], Radhika Iyer[1,2], Khushboo Sheth[1,2], Orr Sharpe[1,2], Navin L. Rao[4], Stephane Becart[5], Julie A. Carman[4], Eddie A. James[6], Jane H. Buckner[6], Kevin D. Deane[7], V. Michael Holers[7], Susan M. Goodman[8,9], Laura T. Donlin[8,9], Mark M. Davis[3,10], William H. Robinson*[1,2,3]

**Affiliations:**

[1]Division of Immunology and Rheumatology, Stanford University, Stanford, CA 94305, USA

[2]VA Palo Alto Health Care System, Palo Alto, CA, 94304, USA

[3]Institute for Immunity, Transplantation and Infection, Stanford University, Stanford, CA, USA

[4]Immunology Discovery, Janssen Research and Development LLC, Spring House, PA 19477, USA

[5]Immunology Discovery, Janssen Research and Development LLC, San Diego, CA 92121, USA

[6]Benaroya Research Institute, Seattle, WA 98101, USA

[7]Division of Rheumatology, University of Colorado School of Medicine, Aurora, CO 80220, USA

[8]Hospital for Special Surgery, New York, NY 10021, USA

[9]Weill Cornell Medicine, New York, NY 10021, USA

[10]Department of Microbiology and Immunology, Stanford University, Stanford, CA 94305, USA

[#]These authors contributed equally: Jae-Seung Moon, Shady Younis

**Supplementary Table 1. Demographic characteristics of RA patients.**

| | **RA patients** |
|---|---|
| Total Number | 59 |
| Gender, Female/Male | 4/55 |
| Age (years) - mean±SD | 66.15±12.24 |
| Dis duration (years) - mean±SD | 11.35±10.69 |
| Disease activity | |
| ACPA positive (%) | 76.27 |
| RF positive (%) | 73.33 |
| CDAI score - mean±SD | 15.17±15.95 |
| MTX use (%) | 51.11 |
| MTX duration (years) | 4.96±4.9 |
